# Changes in Muscle Activation and Joint Motion During Walking After Transtibial Amputation with Sensory Feedback from Spinal Cord Stimulation: A Case Study

**DOI:** 10.1101/2024.12.27.24319684

**Authors:** Rohit Bose, Bailey A Petersen, Devapratim Sarma, Beatrice Barra, Ameya C Nanivadekar, Tyler J Madonna, Monica F Liu, Isaiah Levy, Eric R Helm, Vincent J Miele, Lee E Fisher, Douglas J Weber, Ashley N Dalrymple

## Abstract

**Background:** The goal of this study was to examine the effects of spinal cord stimulation (SCS) on muscle activity during walking after lower-limb amputation. Amputation results in a loss of sensory feedback and alterations in gait biomechanics, including co-contractions of antagonist muscles about the knee and ankle, and reduced pelvic obliquity range-of-motion and pelvic drop. SCS can restore sensation in the missing limb, but its effects on muscle activation and gait biomechanics have not been studied in people with lower-limb amputation.

**Methods:** This case study included a participant with transtibial amputation who was implanted percutaneously with SCS electrodes over the lumbosacral enlargement for 84 days. SCS was used during in-lab experiments to provide somatosensory feedback from the missing limb, relaying a sense of plantar pressure when the prosthesis was in the stance phase of the gait cycle. We used electromyography (EMG) to record muscle activity from the residual and intact limbs, and 3D motion capture to measure pelvic obliquity. EMG signals were recorded during walking with and without SCS at early and late time points across the implant duration.

**Results:** During walking, co-contraction of knee antagonist muscles was reduced following multiple sessions of SCS-mediated sensory restoration. Additionally, the activation of the hip abductor (tensor fasciae latae) muscle increased activity during gait with SCS-mediated sensory restoration, which corresponded to an increase in pelvic obliquity range-of-motion and pelvic drop, towards normal.

**Conclusions:** Restoring sensation in the missing limb using SCS modulated muscle activity during walking leading to improved coordination and pelvic motion in an individual with lower-limb amputation.

## INTRODUCTION

It is estimated that lower-limb amputation will affect over 3.6 million people in the United States by the end of 2050 [1,2]. Most cases of lower-limb amputation are caused by complications from vascular disease (54%) or trauma (45%) [1,2]. During walking, people with lower-limb amputation have difficulty maintaining postural stability, leading to an increased risk of falls in this population [3–6]. This impaired stability and increased risk of falls is, in part, due to a lack of somatosensory feedback from the missing foot and prosthesis [3,4]. Thus, restoring somatosensory feedback from the missing limb is crucial for reducing the fall risk in people with lower-limb amputation.

Indeed, several research groups have implanted electrodes into or around peripheral nerves in the residual limb to deliver electrical stimulation and evoke sensations in the missing limb [7–10]. Recently, we demonstrated that electrical stimulation of the lateral spinal cord was also effective at evoking sensory percepts in the missing limbs of people with upper– and lower-limb amputations [11–13]. Collectively, these works have demonstrated that providing somatosensory feedback from the missing limb can improve standing and walking, as determined by clinical assessments of balance and gait [7– 9,11]. Somatosensory feedback provided by lateral spinal cord stimulation (SCS), in particular, improved balance and gait stability during the most difficult conditions in the Sensory Organization Test and Functional Gait Assessment [11].

Lateral SCS excites the dorsal spinal roots, which contain sensory afferent axons [14,15]. Excitation of sensory afferents results in reflexive activation of motoneurons. Spinal reflexes evoked by SCS are referred to as posterior root-muscle reflexes and can be recorded using electromyography (EMG) [16– 19]. Spinal reflexes that are engaged by somatosensory feedback play a pivotal role in modulating lower-limb muscle activity during walking [20–22]. For example, somatosensory feedback augments limb loading during the stance phase of the gait cycle, [23,24], can prompt the transition from stance to swing [25], and initiates corrective responses to perturbations during walking [21,26,27]. We have recently shown that SCS for sensory feedback engages spinal reflex pathways [28]. Importantly, we demonstrated that the step cycle duration and limb alternation symmetry, which were initially within expected range, were not altered by SCS-evoked reflexes.

Currently, it is unknown if long-term use of somatosensory feedback provided by SCS modulates muscle activity during walking. In this case study in one participant with a unilateral transtibial amputation, we report a reduction in co-contractions of the knee antagonist muscles following multiple weeks of in-lab use of SCS. Furthermore, we observed an increase in EMG activity during walking in the tensor fasciae latae (TFL) muscle (hip abductor), which corresponded to an increase in pelvic obliquity range-of-motion and pelvic drop.

## METHODS

All procedures were approved by the Institutional Review Board at the University of Pittsburgh and conformed to the Declaration of Helsinki. Experiments were performed under an Investigational Device Exemption from the United States Food and Drug Administration and registered at ClinicalTrials.gov (NCT04547582). Prior to their enrollment in the study, the participant provided written informed consent. This case study arose from a study aiming to restore sensations in the missing limb, improve stability during standing and walking, and to reduce phantom limb pain using SCS [11].

### Participant

One individual with transtibial amputation is included in this case study. This participant was a woman between the ages of 60 and 65 years old. Her left lower leg had been amputated 5 years prior to the study because of complications from diabetic neuropathy and dysvascular disease. She was a limited community ambulator, as determined by the Amputee Mobility Predictor [29]. She regularly used a non-motorized lower-limb prosthesis to walk for approximately 52 months before implant. The implant duration was 84 days.

### Spinal cord stimulation lead implant procedure

Under twilight anesthesia, we implanted SCS leads percutaneously in an outpatient procedure. While the participant was lying prone, we inserted three 8-contact SCS leads (Octrode, Abbott Laboratories, Chicago, IL) into the dorsal epidural space near the T12-L2 vertebral levels (Fig 1B) using a 14-guage 4-inch epidural Tuohy needle. Under live fluoroscopy guidance, the leads were steered posterior-laterally using a stylet to target the caudal lumbosacral enlargement, which includes the dermatomes corresponding to the distal residual limb and missing foot. Using an external stimulator, we intraoperatively delivered electrical stimulation through several electrodes on each SCS lead and iteratively adjusted the lead placement according to the participants’ verbal report of the location of evoked sensations in the residual limb. We anchored the SCS leads to subcutaneous fascia via a small incision. For the first 4 weeks, we monitored lead location and migration using weekly X-rays, then performed X-rays twice monthly thereafter. At the conclusion of the study, we removed all percutaneous leads and closed all incisions.

**Figure 1.**
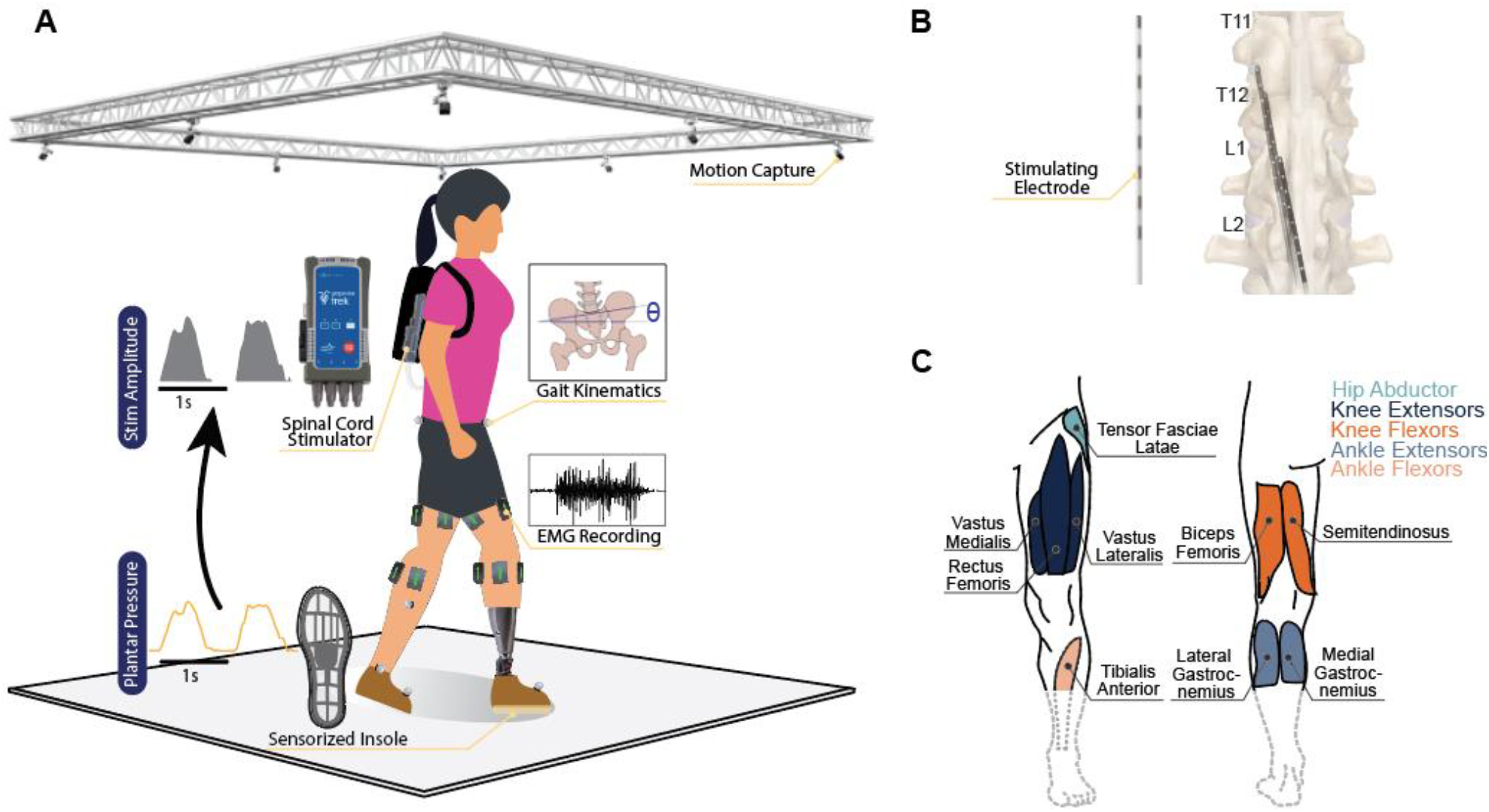
Recording setup and SCS electrode locations. (A) Walking with real-time spinal cord stimulation (SCS). Loading recorded from sensorized insoles linearly modulated the stimulation amplitude. SCS provided sensory feedback. Electromyography (EMG) recorded muscle activity from the residual and intact limb muscles. Motion capture recorded limb position for kinematic analysis. (B) Location of the SCS electrodes, as confirmed from X-rays, relative to spinal vertebral segments at 1-week post-implant. (C) Schematic of the muscles where EMG was recorded. Functional groupings of muscles are indicated by color: hip abductor (teal), knee extensors (dark blue), knee flexors (dark orange), ankle extensors (light blue), and ankle flexors (light orange).

### Spinal cord stimulation protocol

We delivered electrical stimulation pulses using three 32-channel constant current stimulators (Nano 2+Stim; Ripple, Salt Lake City, UT) with a custom circuit board that shorted together groups of four channels to increase the channel amplitude output from 1.5 mA to 6 mA [11,28]. SCS output parameters were controlled using custom software in MATLAB (MathWorks, Natick, MA, USA). Stimulation pulses were cathodic-leading, symmetric and charge-balanced, delivered in a monopolar configuration, with an external return electrode (Natus Disposable Ground Electrode, MVAP Medical Supplies, CA) placed over the right acromion.

### Electromyography recording during walking

We recorded EMG while the participant was walking at a self-selected speed. We prepared the skin overlying the muscles of both legs by cleaning with abrasive gel (Lemon Prep, Mavidon, USA) and alcohol wipes (Nuprep, Weaver, Aurora, CO). We positioned bipolar pairs of surface EMG electrodes (Ag|AgCl disposable dual EMG electrodes, MVAP Medical, Thousand Oaks, CA) on the following proximal leg muscles bilaterally: hip abductor (TFL), knee extensors (rectus femoris (RF) and vastus lateralis (VL)), knee flexors (biceps femoris (BF), and semitendinosus (ST)), ankle flexor (tibialis anterior (TA)), and ankle extensors (lateral gastrocnemius (LG) and medial gastrocnemius (MG)) (Figure 1). The putative distal muscles of the residual limb were located using palpation and guided contraction. We placed a ground electrode (Natus Disposable Ground Electrode, MVAP Medical Supplies, CA) on the anterior superior iliac spine (ASIS) ipsilateral to the amputation. EMG data were collected using a wireless data acquisition system (Trigno Avanti system, Delsys, Natick, MA; sampling rate: 2 kHz; input range: ± 22mV; sensor bandwidth: 10 – 850 Hz).

### Gait kinematics recording

We recorded gait kinematics using a 16-camera OptiTrack motion analysis system (Natural Point, OR, USA) while the participant was walking. We placed 16 reflective markers on anatomical landmarks according to the OptiTrack “Conventional Lower Body” model [30,31]. Motion capture data were sampled at 100 Hz.

### Study session protocol

The participant attended 2-4 testing sessions per week throughout the implant period. Within these sessions, we tested sensory responses to SCS [11], recorded reflex responses [28], and provided sensory feedback during walking. No stimulation was delivered outside of the lab environment.

We delivered SCS during overground walking starting on Day 22 post-implant. We instructed the participant to walk at their self-selected speed across a 6-meter walkway. Wireless plantar pressure sensing insoles (Moticon Insole 3, Munich, Germany) measured limb loading. When the plantar pressure of the insole in the prosthetic foot exceeded a threshold to indicate the stance phase, the stimulation was triggered to evoke sensations in the participant’s missing foot. Therefore, SCS was delivered only during the stance phase of the prosthetic limb. Stimulus amplitude was linearly modulated with insole pressure above a threshold of 3 N/cm^2^. Stimuli were delivered at 90 Hz with a pulse width of 0.2 ms.

### Data analysis methods

#### Quantifying muscle activity during walking

We sought to quantify muscle activation amplitude and timing during walking with stimulation at a self-selected speed compared to walking without stimulation. We extracted the EMG envelope by first applying a high-pass Butterworth filter with a cut-off frequency of 0.1 Hz, then rectifying. We smoothed the EMG envelope using a moving average of the rectified signal, with a bin size of 200 samples. A z-transform was applied to the smoothed EMG envelope to standardize the amplitude for each muscle.

Step cycles were segmented using the TA EMG from the intact limb to detect heel-strike event times. Specifically, we marked 10% of maximal activation of the TA muscle at offset as 20% of the gait cycle, based on previous reports of muscle activation during gait [32,33]. Using that marker, we aligned all EMG data from all muscles accordingly.

To compare the timing and overall activation of the muscle profiles during walking, we first time-normalized each step to align 0 – 100% of the gait cycle. We then found the average and standard deviation of the single-step EMG envelopes. Next, we calculated the cross correlation of the average EMG envelopes across conditions. From the cross correlation, we obtained a measure of similarity and a time lag between the pairs of EMG envelopes. Additionally, we determined the difference in overall muscle activation magnitude by calculating and comparing the area-under-the-curve of the average EMG envelope.

In order to quantify the co-contraction of antagonist muscles across the knee joint during walking, we calculated the co-contraction area using the integrated activation of the EMG envelopes from the VL and BF muscles across conditions [34,35]:

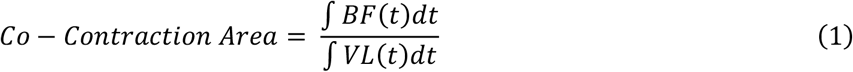

where a larger co-contraction area indicates greater co-contraction of these antagonist muscles. We compared the co-contraction area at the early and late time points with and without SCS using a one-way analysis of variance (ANOVA) and Bonferroni post-hoc tests.

Due to an error with some wireless sensors during data collection, we do not have EMG data for all muscles during walking. Therefore, we were unable to assess co-contractions of the ankle muscles, nor were we able to assess changes in TFL activity in the intact limb.

#### Gait kinematics

We sought to characterize how muscle activation and timing induced changes in gait kinematics during walking at a self-selected speed with SCS as compared to walking without SCS. We also characterized the effect of multiple exposures to SCS over the study duration by comparing muscle activation during walking without stimulation at the early and later time points of the implant. Because we observed changes in TFL muscle activity (described in detail below), we measured the pelvic obliquity during walking. Pelvic obliquity is the rotation of the pelvis in the coronal-plane (Figure 5B). We filtered the marker positions over time using a fourth order low-pass Butterworth filter (cut-off frequency = 20 Hz). We calculated the pelvic obliquity (θ) during walking using the left and right ASIS motion capture marker 3-dimensional positions [36]:

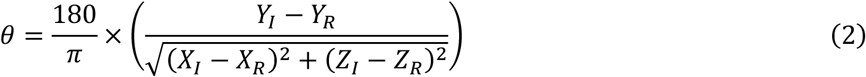

where the *X, Y*, and *Z* coordinates correspond to the medial-lateral, rostral-caudal, and anterior-posterior dimensions, respectively, and *I* and *R* are the intact and residual limb, respectively. When the ASIS markers were equal in elevation, the pelvic obliquity was 0º. When the ASIS marker on the residual limb was tilted higher than the intact limb, the pelvic obliquity was negative. From the pelvic obliquity, we extracted two measures: the range-of-motion and pelvic drop. The range-of-motion is the maximal peak-to-peak value of the pelvic obliquity for a single step (Figure 5A). The pelvic drop is the downward rotation angle of the contralateral hip from the time of ipsilateral foot contact to the maximum pelvic obliquity [36]. We measured the pelvic drop for both the intact and residual limbs.

We compared the pelvic obliquity range-of-motion and pelvic drop across all three time points, and compared the pelvic drop between the intact and residual limbs. To compare the pelvic obliquity range-of-motion and pelvic drop across conditions, we used a permutation test, which is a non-parametric statistical approach often used for smaller sample sizes [37]. The permutation test entails shuffling the observed data with replacement, maintaining the number of measurements in each condition. We obtained 10,000 resampled and shuffled sets of data, and for each set, calculated the difference in means. We compared the difference in means from each of the permutations of the resampled data with the difference in means of the observed data. We obtained a one-sided p-value by calculating the proportion of resampled permutations where the difference in means was greater than the observed mean, and dividing that by the number of repetitions (10,000). We used the Shapiro–Wilk test to test for normality and Levene’s test to assess the homogeneity of variance. For all statistical analyses, we considered a p-value < 0.05 to indicate significance.

## RESULTS

### Hip abductors were more active following multiple weeks of SCS

We compared the activation of muscles during walking without sensory stimulation at Days 30 and 63 post-implant, and with and without stimulation at Day 63 (testing of walking with sensory feedback began on Day 22 post-implant). The step cycle duration with and without SCS were equivalent within ± 10% [28].

We generated EMG envelope profiles of muscles across the residual and intact limbs throughout the gait cycle without stimulation over time and at the later time point with stimulation. Overall, the muscle activation profiles with and without SCS at the later time point were similar across all muscles, exhibiting high cross-correlation values (all > 90%; Figure 2A). The residual TFL, intact LG, and intact TA had zero lag between the envelopes, whereas the residual VL, residual BF, and intact MG exhibited small changes in timing: the envelope from the with-SCS condition was shifted earlier by < 6% of the gait cycle compared to without SCS. Without SCS at the early and later time points, the residual TFL muscle had moderate cross correlation equal to 72.9% and the rest of the muscles strongly correlated (> 96%; Figure 3A). The residual TFL and VL muscles were more active at the later time point compared to the earlier time point, with 77.6% and 81.7% increases in amplitude, respectively (Figure 3B). The TFL muscle was 93.6% more active at the later time point with SCS compared to the early time point without SCS (Figure 2B, 3C). With SCS, the other muscles exhibited small changes in amplitude that were less than 8%.

**Figure 2.**
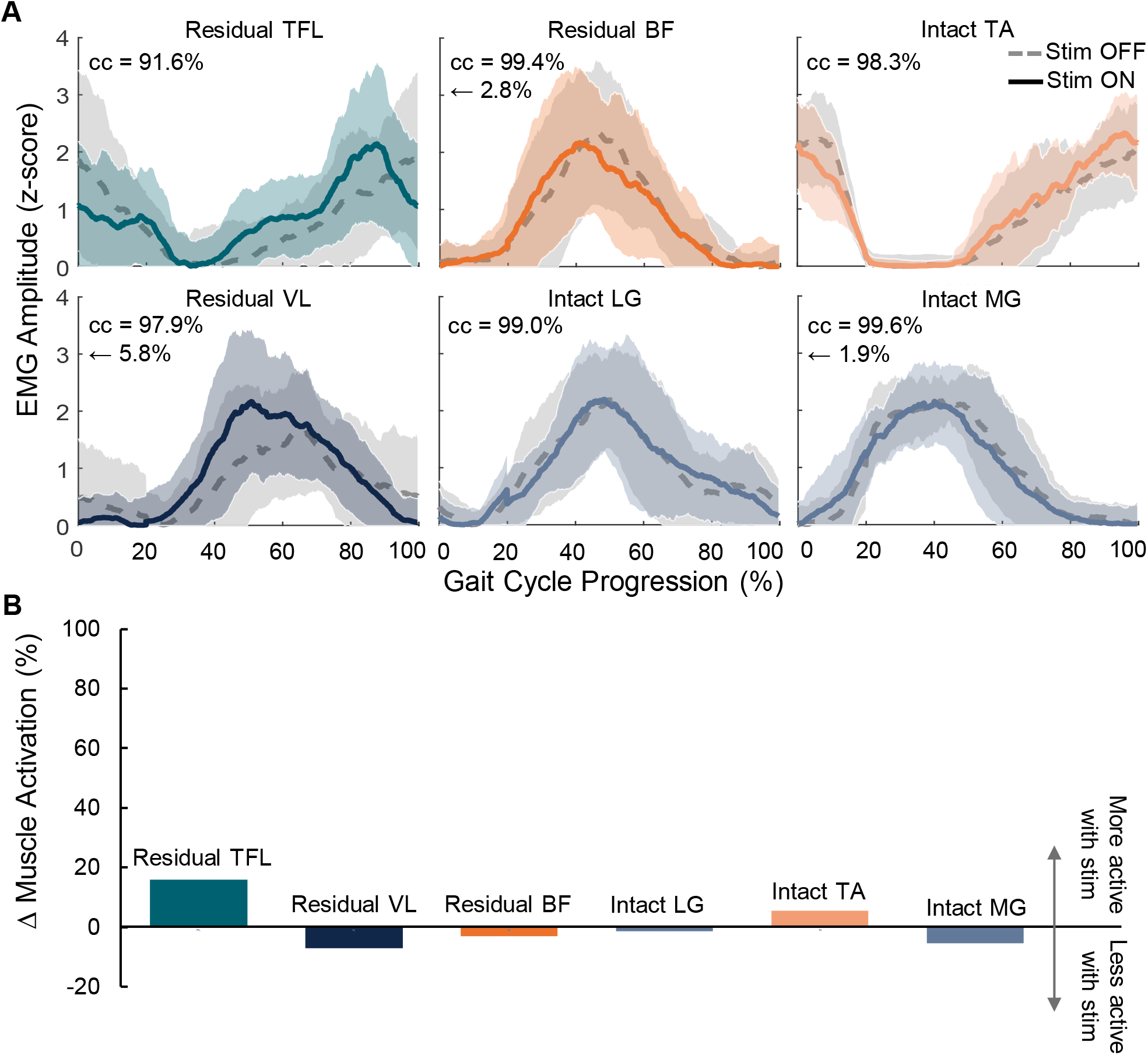
Muscle activity during walking with and without spinal cord stimulation (SCS) on Day 63. (A) Mean ± standard deviation (SD) EMG envelopes throughout the gait cycle, with (solid line) and without (dashed line) SCS. Stim = stimulation; cc = cross correlation of the two EMG envelopes; arrow indicates relative shift of EMG envelope with SCS compared to without, expressed as a percentage of gait cycle. TFL = tensor fasciae latae; BF = biceps femoris; LG = lateral gastrocnemius; TA = tibialis anterior; MG = medial gastrocnemius. (B) Change (Δ) in muscle activation amplitude. Positive change = muscle was more active with SCS than without.

**Figure 3.**
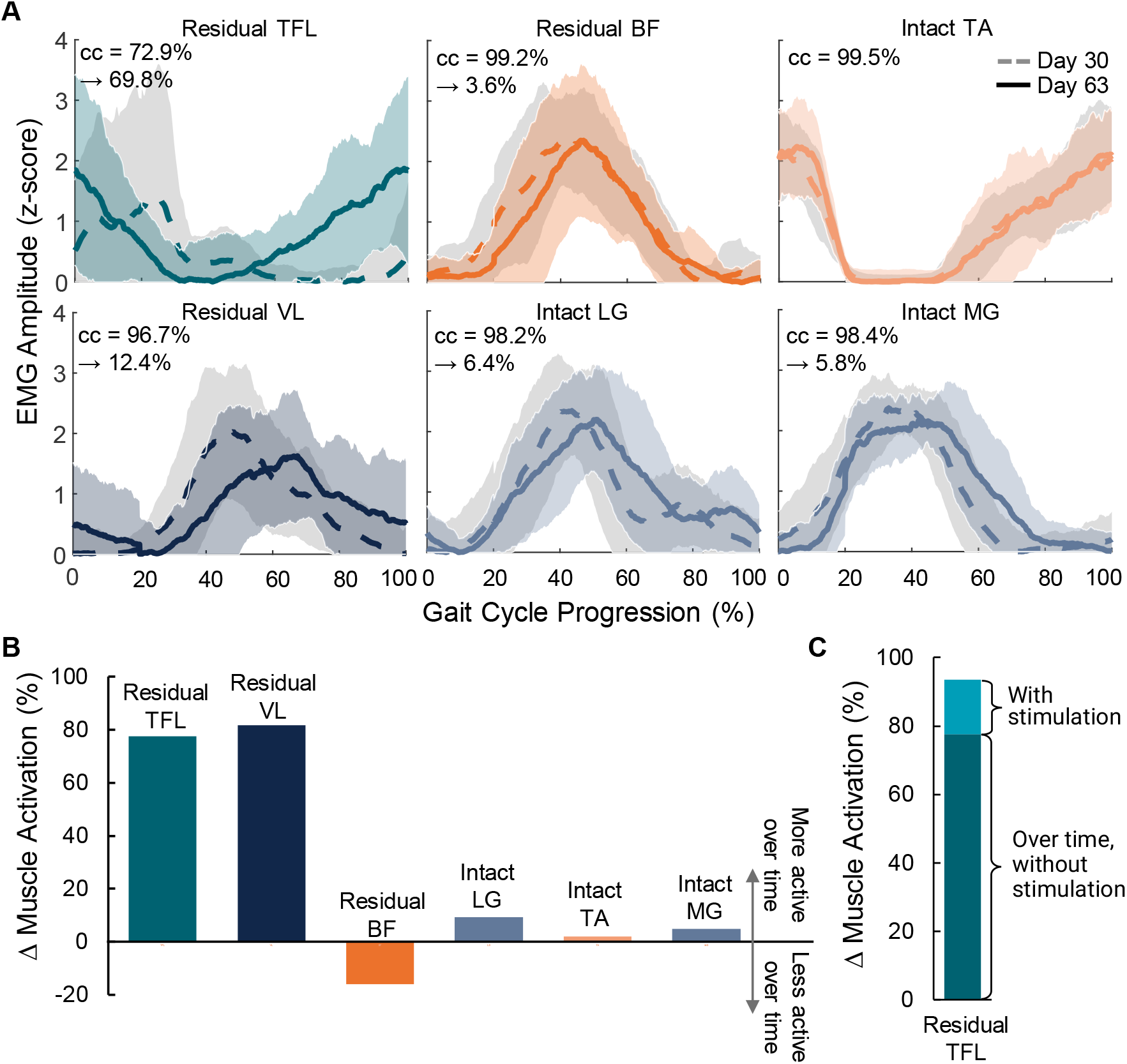
Muscle activity during walking without spinal cord stimulation (SCS) on Days 30 and 63. (A) Mean ± standard deviation (SD) EMG envelopes throughout the gait cycle, with (solid line) and without (dashed line) SCS. cc = cross correlation of the two EMG envelopes; arrow indicates relative shift of EMG envelope with SCS compared to without, expressed as a percentage of gait cycle. TFL = tensor fasciae latae; BF = biceps femoris; LG = lateral gastrocnemius; TA = tibialis anterior; MG = medial gastrocnemius. (B) Change (Δ) in muscle activation amplitude. Positive change = muscle was more active with SCS than without. (C) Change in muscle activation of TFL over time without and with SCS.

### Co-contractions of knee antagonist muscles were reduced following multiple weeks of SCS

At the early time point without SCS, there was a high co-contraction area between the VL and BF muscles of the residual limb during walking (co-contraction area = 1.57 ± 0.52; Figure 4). At the later time point, the co-contraction area reduced significantly to 1.05 (±0.35; p < 0.001) without stimulation and 0.95 (± 0.33; p < 0.001) with stimulation, meaning that there was a reduced co-contraction between the knee antagonist muscles of the residual limb over time, with a further, but slight, reduction in co-contraction when SCS was turned on.

**Figure 4.**
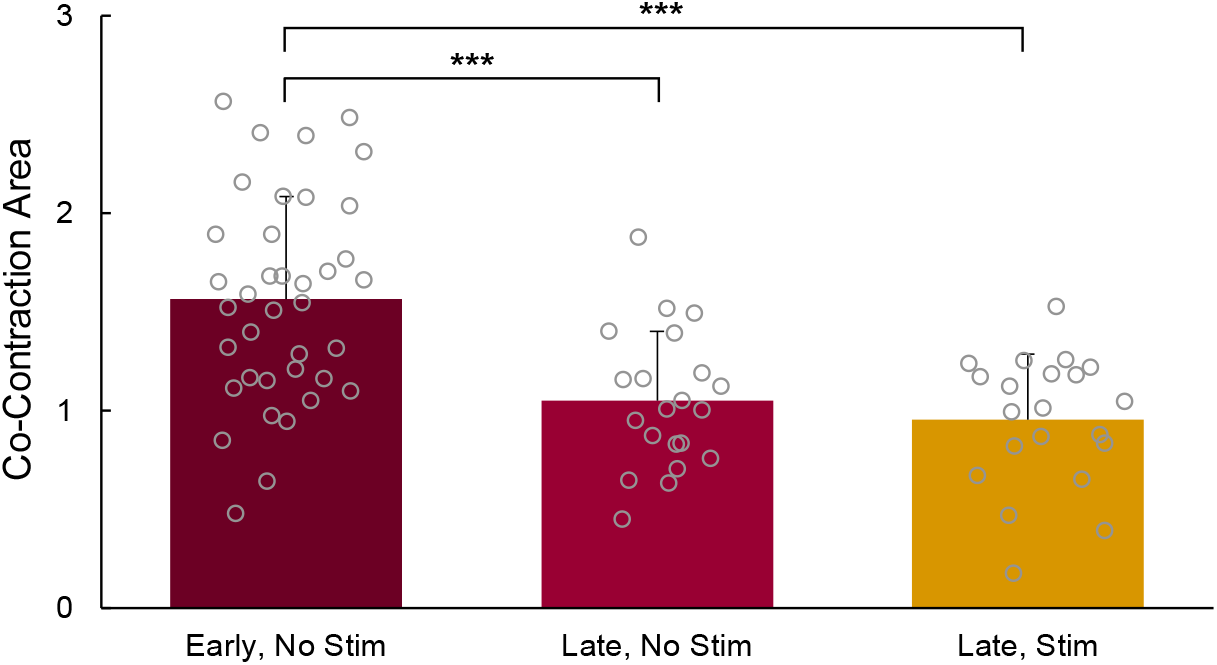
Mean (+ SD) of the co-contraction area, indicating co-contractions of the biceps femoris and vastus lateralis muscles, at the early and late timepoints with and without SCS. ***p < 0.001.

**Figure 5.**
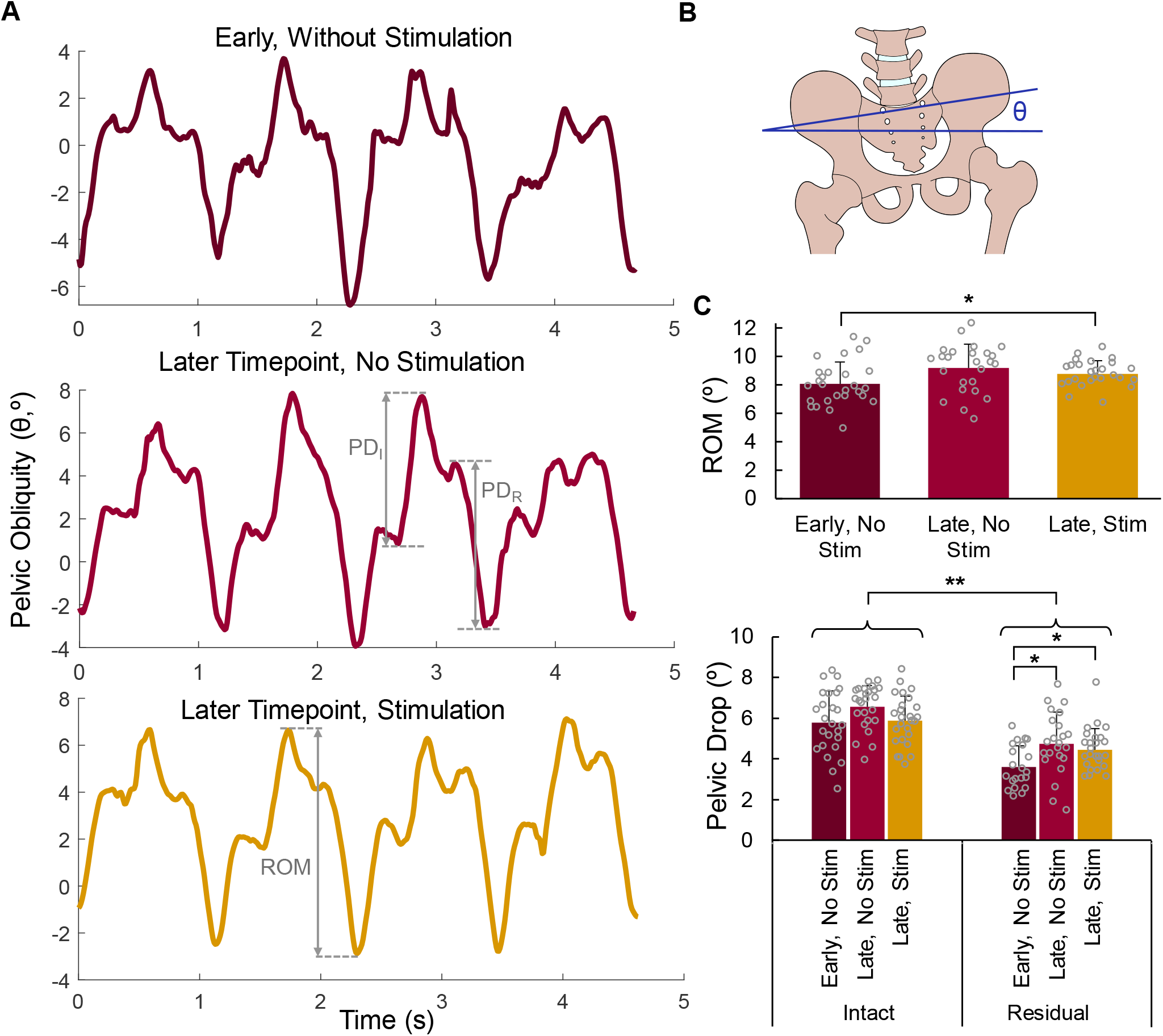
Pelvic obliquity during walking. (A) Representative pelvic obliquity (θ) at early (Day 30) and late (Day 63) time points, without and with spinal cord stimulation (SCS). Pelvic drop (PD) is the downward rotation angle of the contralateral hip from the time of ipsilateral foot contact to the maximum pelvic obliquity. Range-of-motion (ROM) is the peak-to-peak amplitude of the pelvic obliquity for a single step. I = intact limb, R = residual limb. (B) Illustration of pelvic obliquity. (C) ROM (top) and PD (bottom) at the early and later time points without SCS, and the later time point with SCS. PD includes differences between the intact and residual limbs. Stim = stimulation. *p < 0.05; **p < 0.01.

### Pelvic obliquity range-of-motion and pelvic drop increase following multiple weeks of SCS

The TFL muscle plays a role in pelvic obliquity. Because TFL activity of the residual limb showed the largest changes after multiple weeks of SCS, we characterized the functional outcomes of this change in activation. Using the motion capture data, we measured the pelvic obliquity during walking (Figure 5A). The pelvic obliquity range-of-motion at the early time point without SCS was 8.08º (± 1.55º) (Figure 5C). At the later time points, the range-of-motion increased to 9.20º (± 1.69º) and 8.80º (± 0.91º), without and with SCS, respectively. The increase in range-of-motion at the later time point with SCS was significantly different from the early time point without SCS (p = 0.027). The range-of-motion at the later time point without SCS was not different from the early time point without SCS (p = 0.065) nor the later time point with SCS (p = 0.212). Overall, the pelvic drop of the residual limb was significantly smaller than the intact limb (p = 0.002; Figure 5C). The pelvic drop of the residual limb increased significantly from 3.62º (± 1.04º) at the early time point to 4.76º (± 1.53º) at the later time point without SCS (p = 0.022). Furthermore, the pelvic drop of the residual limb was significantly higher at the later time point with SCS (4.46º ± 1.04º) compared to the early time point without SCS (p = 0.034). The pelvic drop at the later time point with and without SCS were not significantly different (p = 0.258). Therefore, the increase in TFL activity with SCS corresponded to functional improvements in pelvic obliquity range-of-motion and pelvic drop.

## DISCUSSION

Our previous work showed that SCS can be used to evoke sensory percepts in a missing limb [11–13]. Reflexes evoked by SCS did not interfere with normal measures of gait including gait cycle duration and limb alternation symmetry [28]. Activation of spinal reflex pathways using SCS can elicit changes in muscle activation following neural injury [38–40]. The primary goal of this study was to quantify changes in muscle activation in a participant with a transtibial amputation who received SCS to restore sensation in their missing limb. Changes in muscle activity following extended time using SCS in the lab corresponded with functional improvements in hip motion and reduced co-contractions of knee antagonist muscles.

### Restoring sensation using SCS improved joint stability and range of motion during walking

People who use a leg prosthesis exhibit multiple gait abnormalities arising in part from the loss of sensation from the missing limb, reducing their confidence in weight-bearing on the prosthetic limb [3– 6]. As a result, prosthesis users will employ compensatory movements to increase their stability and confidence as they walk. For example, knee flexors and extensors of the residual limb co-contract more than the intact limb during walking in people with transtibial amputations [35]. Co-contraction of antagonist muscles about the knee is a compensatory strategy to increase stiffness in preparation for foot-contact and weight-bearing. Prosthesis users will also exhibit exaggerated hip hiking (raised hip) or circumduction (abduction and semi-circular movement) of the prosthetic limb during the swing phase to ensure that the foot clears the ground [36]. These compensatory hip movements stem from insufficient residual limb knee bending and lack of ankle dorsiflexion during swing, resulting in toe dragging, which could cause tripping. Pelvic obliquity is a measure of pelvic rotation in the coronal-plane and can be used to indicate hip hiking [36]. Specifically, during the stance phase of walking, hip abductors on the stance leg contract to tilt the pelvis and raise the contralateral hip to aid with foot clearance, resulting in an increased pelvic obliquity. At heel strike, there are small changes in pelvic obliquity, suggestive of improved shock absorption [41]. People with a lower-limb amputation exhibit a reduced pelvic obliquity range-of-motion during gait, with a further decrease in range-of-motion for higher-level amputations [36]. Furthermore, the pelvic drop of the stance limb is smaller on the prosthetic limb than the intact limb [36]. In this study, at the later time point with and without SCS, the pelvic drop of the residual limb became more symmetrical with the intact limb. Additionally, at the later time point with and without SCS, the pelvic drop of the residual limb increased towards values seen in neurologically-intact adults (5 – 7°) [36].

We demonstrated in other work that SCS to restore sensation activates spinal sensorimotor reflex pathways during walking [28]. It is possible that, by activating these reflex pathways, over time, SCS modulates muscle activity to provide additional and necessary joint stabilization during walking [20–22]. SCS was delivered only during the stance phase of the prosthetic limb, and we found the most drastic change in VL and TFL activity during this phase. The VL muscle is a knee extensor and provides stability to the knee joint during the stance phase. At the later time point in the study, the residual VL muscle was more active than the early time point. Furthermore, the residual BF muscle was slightly less active at the later time point in the study compared to the earlier time point in the study. The increase in VL activity and decrease in BF activity reflect the reduced co-contraction area of the residual limb during walking at the later time point. Therefore, instead of compensatory co-contraction of the antagonist muscles to provide stability to the knee joint, knee stability was driven primarily by the VL muscle following SCS. The TFL muscle is a hip abductor; during the stance phase, the TFL muscle contracts, dropping the ipsilateral pelvis, allowing the contralateral pelvis to raise and the contralateral foot to clear the ground [42]. Our results at the early time point align with the previous study showing a decrease in pelvic obliquity range-of-motion and pelvic drop of the residual limb [36]. At the later time point, the increased activation of the residual TFL muscle during the stance phase with and without SCS is indicated by the increased pelvic drop and pelvic obliquity range-of-motion. Therefore, we demonstrate in this case study that following repeated exposure to SCS-mediated sensory restoration, there are functional improvements in dynamic stability at the knee and the hip during walking. The functional improvements in pelvic drop were present at the later time point, even without stimulation, suggesting a possible rehabilitative effect of SCS on muscle activation and knee and hip motion during walking, but data from additional participants is needed to be conclusive. This study is the first to investigate changes in muscle activity alongside the functional outcomes following long-term use of a sensory neuroprosthesis.

### Limitations

During the first few weeks of the study, much of the time was spent characterizing the sensory percepts in the missing limb; therefore, more extensive testing with SCS for sensory feedback was only possible with this single participant who had a longer implant duration (compared to other participants that were implanted for only 28 days [11,28]). Nonetheless, we demonstrate reduced co-contractions of knee antagonist muscles and improved pelvic obliquity range-of-motion and pelvic drop following extended, in-lab delivery of SCS. Future studies will build on the current case study to further characterize the functional improvements and corresponding changes in muscle activity during walking with long-term SCS.

### Clinical relevance

SCS is a common method to reduce pain, with as many as 50,000 people receiving a SCS implant per year to treat chronic pain [40,43]. Through our previous work and the current study, we demonstrate that, in addition to reducing phantom limb pain, SCS can be used to restore sensation in the missing limb, improve function during walking [11], and stabilize the residual limb, particularly the knee and pelvis, during walking. Therefore, with improved sensorimotor function, over time, SCS may be able to correct abnormal torque on residual and intact limb joints, resulting in less long-term pathological consequences of compensatory movements, such as arthritis. Additionally, SCS may reduce the incidence of falls in people with lower limb amputation. Future studies will focus on additional gait tasks to further characterize the role of muscle activation on changes in gait over time with sensory feedback.

## CONCLUSIONS

SCS was used to restore sensation in the missing limb in a person with a transtibial amputation during walking. Sensory feedback from SCS resulted in changes in muscle activation in the residual limb, leading to functional improvements during walking. These changes included reduced co-contractions of the knee antagonist muscles and increased activation of the TFL muscle, corresponding to increased pelvic obliquity range-of-motion and pelvic drop.

## Data Availability

All data produced in the present study are available upon reasonable request to the authors

## LIST OF ABBREVIATIONS

SCS: spinal cord stimulation
EMG: electromyography
TFL: tensor fasciae latae
RF: rectus femoris
VM: vastus medialis
VL: vastus lateralis
BF: biceps femoris
ST: semitendinosus
TA: tibialis anterior
LG: lateral gastrocnemius
MG: medial gastrocnemius
SD: standard deviation
ANOVA: analysis of variance
ASIS: anterior superior iliac spine

## ACKNOWLEDGMENTS

The authors would like to express our sincerest gratitude to our research participants for their time and dedication to furthering the field of neuroprosthetics. We would like to thank Debbie Harrington, Casey Konopisos, and Alayna Schwerer for their assistance with the IRB protocol, clinical trial registration, IDE application, and participant recruitment.

## FUNDING SOURCE

This study was funded by the National Institutes of Health (NINDS Award number UH3NS100541 and NICHD Award Number F30HD0987984).

## CLINICAL TRIAL INFORMATION

This study was registered under clinical trial NCT04547582.

## COMPETING INTERESTS

DJW is a founder and shareholder of Reach Neuro, Inc.; DJW is a consultant and shareholder of Neuronoff, Inc. and Panther Life Sciences, Inc.; DJW is a shareholder and scientific board member for NeuroOne Medical, Inc.; DJW is a co-founder and shareholder of Bionic Power Inc. BB is the inventor of several patents involving technologies for the electrical stimulation of the spinal cord. The other authors declare no conflicts of interests in relation to this work.

## Notes

### Clinical Trial

NCT04547582

### Author Declarations

All procedures were approved by the Institutional Review Board at the University of Pittsburgh and conformed to the Declaration of Helsinki.

